# Subclinical perioperative Myocardial Injury following Complex Endovascular Aortic Repair is associated with increased mid-term mortality; a retrospective study

**DOI:** 10.1101/2025.11.20.25340709

**Authors:** DE Vecht, A Vanmaele, V Rastogi, NM Van Mieghem, SE Hoeks, F van Lier, HJM Verhagen, JL de Bruin

## Abstract

**Background:** Perioperative myocardial injury (PMI), defined by an asymptomatic increase in troponin levels, has been recognized as prognostic marker for impaired survival following non-cardiac surgery with therapy for improvement potentially available. Patients undergoing complex endovascular aneurysm repair (c-EVAR) may be particularly at risk for PMI due to their cardiovascular burden. However, due to lacking routine perioperative troponin monitoring, the incidence and impact of PMI on long term mortality following c-EVAR remains unclear with no possibility of applying improvement strategies.

**Methods:** Preoperative and consecutive postoperative high-sensitivity troponin-T (hsTnT) levels were retrospectively analysed from all patients who underwent elective c-EVAR in a Dutch tertiary hospital between 2012 and 2022. PMI was defined as a difference between pre- to postoperative troponin concentrations (ΔhsTnT) of more than ≥14 ng/L in the absence of clinical features of myocardial infarction. Primary objectives were the incidence of PMI following c-EVAR and its association with four-year mortality.

**Results:** 180 patients were included. Median follow-up time was 41 months. 38 patients (21.1%) developed PMI. Coronary artery disease was more prevalent (CAD)(68.4 % vs. 42.3%, p=.012) and ASA-scores were higher in patients with PMI (ASA-4: 13.1% vs. 7.8%, p=.044). PMI was independently associated with increased four-year mortality (adjusted HR: 2.37 (95% CI: 1.25-4.48), p=.008). CAD (adjusted OR: 2.38 (95% CI: 1.07-5.51), p=.037) and preoperative elevated troponin levels (adjusted HR: 2.65 (95% CI: 1.19-6.3, p=.023) were independently associated with an increased risk for PMI.

**Conclusion:** Asymptomatic PMI is relatively common following c-EVAR and is associated with impaired mid-term survival. Given the high prevalence of cardiovascular comorbidities in patients with PMI, prospective follow-up studies should focus on the potential of routine perioperative hsTnT monitoring in identifying high-risk patients who could potentially benefit from enhanced cardiac care.

## Introduction

Perioperative myocardial injury (PMI), defined as an acute rise in troponin levels, but without any clinical symptoms of myocardial ischemia, has emerged as a significant prognostic marker for postoperative impaired survival following non-cardiac surgery, as highlighted in the latest Universal Definition of Myocardial Infarction (4^th^ UDMI).(1) Unlike myocardial infarction, which entails both troponin elevation and clinical evidence of myocardial ischemia, PMI often remains undetected due to its asymptomatic nature (no evident angina and/or ECG-changes) and limited use of routine troponin surveillance in non-cardiac surgery.(2) As a result, the true incidence and prognostic implications of PMI, particularly in high-risk patients, remains therefore uncertain.

Patients undergoing complex endovascular aortic repair (c-EVAR) procedures, a minimally invasive treatment for aortic aneurysms involving the renovisceral arteries, may be particularly vulnerable to PMI. c-EVAR is increasingly used in patients deemed unfit for open surgery, and cardiovascular comorbidities are highly prevalent in these patients.(3,4) Correspondingly, up to 80% of the patients undergoing c-EVAR have cardiovascular comorbidities, highlighting their vulnerability to perioperative cardiac events.(5)

Although the precise pathophysiology remains to be clarified, PMI is thought to arise from an imbalance between cardiac oxygen supply and demand, potentially triggered by perioperative hemodynamic stress and exacerbated by factors such as existing coronary artery disease (CAD), heart failure or anemia.(1,6,7) Furthermore, EVAR itself may impose additional cardiac strain. While endovascular techniques obviate aortic cross-clamping, they do impose their own hemodynamic stress. Experimental work suggests that long stent grafts used in EVAR reduce aortic compliance and consequently increase cardiac afterload, potentially prolonging cardiac strain into the postoperative phase.(8)

Despite this plausible risk, postoperative cardiac surveillance strategies following c-EVAR are poorly defined. It remains unclear to what extent PMI occurs in this population and how it affects postoperative outcomes. Prompt identification of PMI may allow for targeted postoperative evaluation and intervention, with the potential to improve long-term prognosis.(9)

This study aims to investigate the incidence of PMI following c-EVAR, as well as its association with mid-term mortality. Furthermore, the influence of potential risk factors for the development of PMI will be assessed. A better understanding of PMI in this high-risk population could refine risk stratification and guide postoperative cardiac surveillance strategies.

## Methods

### Design and study population

This retrospective study included all patients who underwent c-EVAR for non-ruptured, complex aortic aneurysms (para-, juxta-, supra-renal, or thoracoabdominal), or for inadequate exclusion of infrarenal aneurysms after standard EVAR at a tertiary center (Erasmus University Medical Center, Rotterdam, the Netherlands) between 2012 and 2022. Patients with missing preoperative or postoperative high-sensitivity troponin-T levels (hsTnT) and patients who suffered from AMI within 72 hours after surgery were excluded. The study was approved by the hospital’s medical research ethics committee (METC, MEC-2022-0591) and performed following the guidelines of the Declaration of Helsinki and in accordance with the STROBE-guidelines.

### Variable definitions and data collection

Data on patient demographics, clinical characteristics, medication, procedural and admission details and laboratory reports were obtained from institutional electronic patient records, according to the reporting standards of the Society for Vascular Surgery for complex abdominal aortic repair.(10) c-EVAR stent-grafts include the renovisceral arteries through fenestration (FEVAR) or branches (BEVAR) in the main body. Endografts incorporating both fenestrations and branches were categorized as BEVAR. Smoking status was categorized as non-smoker, smoker, or former smoker when abstained from smoking for at least one month. The estimated glomerular filtration rate (eGFR) was calculated using the Chronic Kidney Disease Epidemiology Collaboration equation (CKD-EPI).(11) Preoperative renal function was categorized as eGFR <60 mL/min or eGFR >60 ml/min, with renal dysfunction defined as a eGFR < 60 mL/min. End stage chronic kidney stage (CKD) was defined as a preoperative eGFR <15 ml/min. Coronary artery disease (CAD) was linked to patients who had a history of myocardial infarction, symptomatic angina pectoris, a positive (CT)-CAD or positive ECG for CAD or a history of coronary revascularization interventions. Included medication involved statin-, beta blocker-, anti-coagulation- and anti-platelet therapy. Anti-platelet therapy was defined as either aspirin, P2Y12-inhibitors or a combination of the two (dual antiplatelet therapy (DAPT)). Mortality data were collected from the Dutch national Personal Records Database on the 9th of September 2024. For the current analyses, data acquisition was stopped on September 2024.

### Troponin measurement and perioperative myocardial injury

Myocardial injury was defined according to the 4^th^ Universal Definition of Myocardial Infarction (4^th^ UDMI), formulated by the European Society of Cardiology (ESC), which requires the diagnosis of myocardial injury to be confirmed by one or more troponin samples above the 99^th^ percentile upper reference limit.(1,2) Preoperative cardiovascular screening followed ESC/European Society of Anesthesiology (ESA) guidelines on high-risk surgery and included collection of cardiac biomarkers (hsTnT and NTproBNP), evaluation of medical history, clinical symptoms, functional capacity (METs) and echocardiography. Coronary angiography ((CT)-CAD) was performed in patients with a history of valvular heart disease or coronary revascularization within six months before undergoing c-EVAR.

HsTnT-levels were measured using the Elecsys assay (F. Hoffmann-La Roche AG, Basel, Switzerland), with the upper 99^th^ percentile set at 14 ng/L.(6) Troponin levels were routinely measured the day before surgery and thrice a week postoperatively, starting on the first postoperative day. The highest available postoperative HsnT-sample within 72 hours after surgery was selected for this study’s analyses. While there is no uniform definition of PMI, a minimum absolute change of at least 14 ng/L from pre- to postoperative maximum hsTnT levels (ΔhsTnT), within 72 hours after surgery was applied (Figure 1).(2,7) In line with the 4^th^ UDMI, acute myocardial infarction (AMI) was defined as a ΔhsTnT ≥14ng/L together with clinical features of myocardial ischaemia such as angina pectoris or a positive ECG or CAG within 72 hours after surgery.(1)

**Figure 1:**
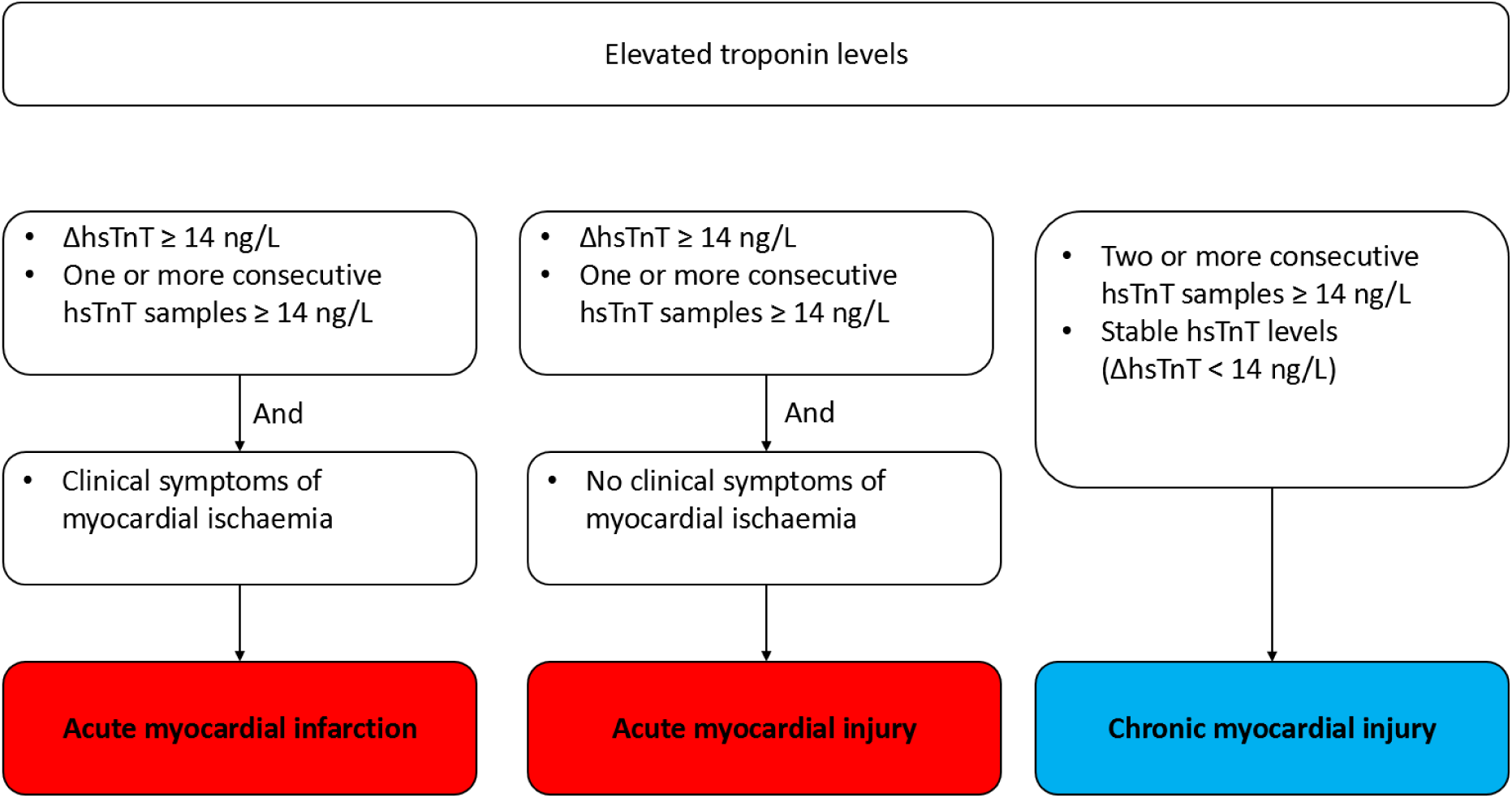
Definitions of myocardial injury and infarction according to the 4^th^ UDMI

### Study objectives

Main study objectives were the incidence of PMI and its association with mid-term all-cause mortality following c-EVAR. Secondary endpoints were the influence of preoperative elevated troponin levels and other cardiovascular variables on the development of PMI.

### Statistical methods

Continuous variables were presented as mean and standard deviation or median and interquartile range, depending on normality of their distribution as assessed visually and confirmed by a Shapiro-Wilk test.

Categorical variables were summarized as counts and percentages. Differences in patient characteristics between patients with and without PMI were tested with independent Student’s t-, Mann Whitney U, Pearson’s chi-squared, or Fisher’s exact tests, as appropriate.

Kaplan-Meier curves and a log-rank test were used to compare four-year mortality between patients with and without PMI after c-EVAR. The risk of mortality associated with PMI was investigated using three consecutive Cox proportional hazards models, first univariable, next, adjusting for age and sex, and finally, further adjusting for sex, age, coronary arterial disease (CAD), renal dysfunction, preoperative elevated hsTnT-levels and ASA score. Proportional hazard assumptions were tested with Schoenfeld individual tests. The relationship between individual preoperative hsTnT levels and probability to develop PMI was visualized using locally estimated scatterplot smoothing (LOESS). Univariate and piecewise analyses were subsequently performed to assess the association between individual preoperative hsTnT-levels with PMI. Multivariable logistic regression was used to evaluate potential risk factors for the development of PMI following c-EVAR.

Statistical analyses were conducted using R version 4.4.0. and higher. All tests were two-tailed, with a significance level of .05.

## Results

Of 213 patients who underwent c-EVAR between 2012 and 2022, 180 patients were used for analysis after excluding 33 patients with either missing hsTnT data or a confirmed AMI within 72 hours postoperatively (Figure 2). Preoperatively, three patients underwent additional screening (CT coronary angiography); two patients because of a recent PCI and one due to a recent episode of angina pectoris. Coronary atherosclerosis was confirmed in these three patients.

**Figure 2:**
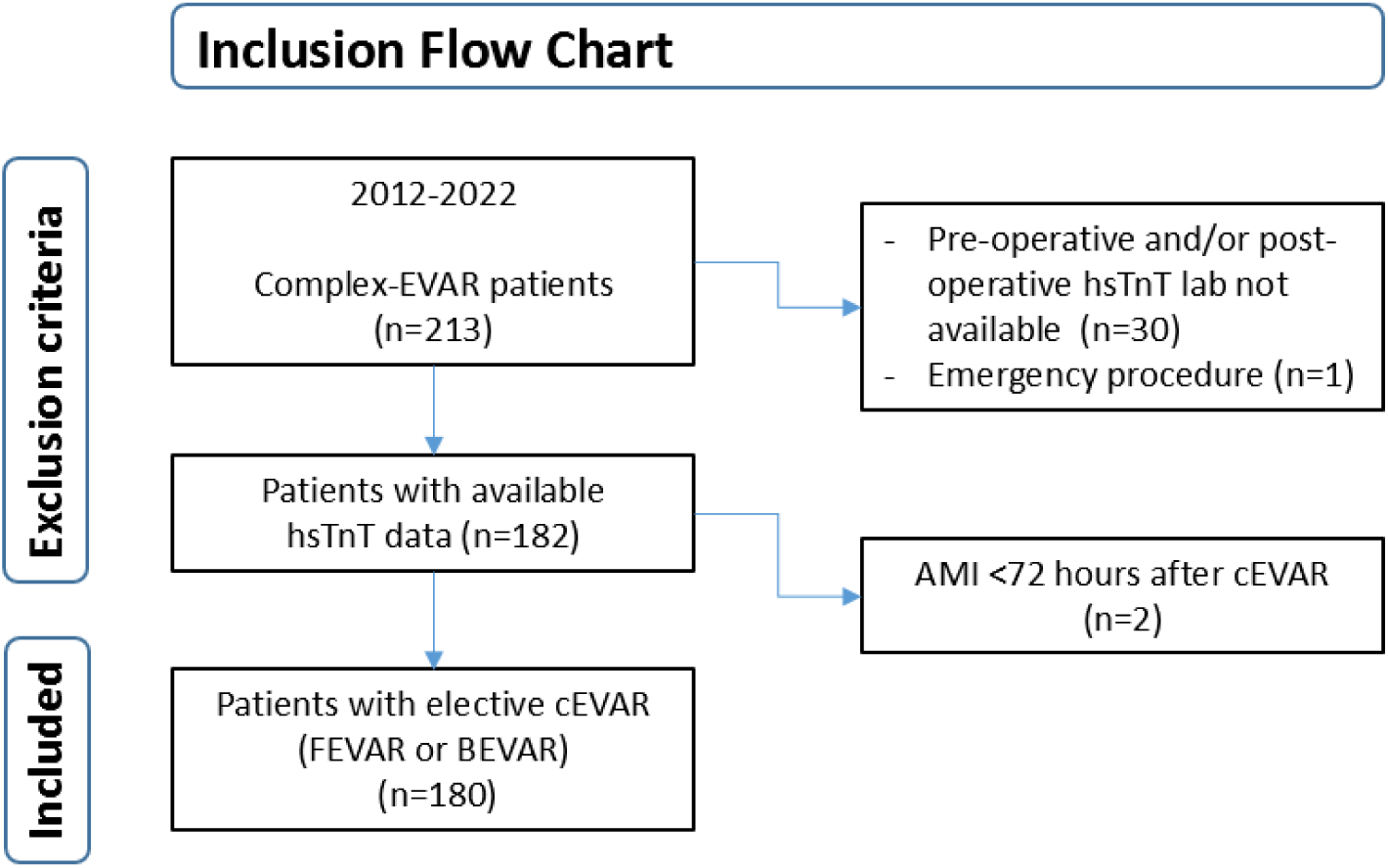
Flow chart showing inclusion of eligible patients; AMI = acute myocardial infarction; F/BEVAR = fenestrated/branched endovascular aneurysm repair; hsTnT = high sensitivity troponin T

**Figure 3:**
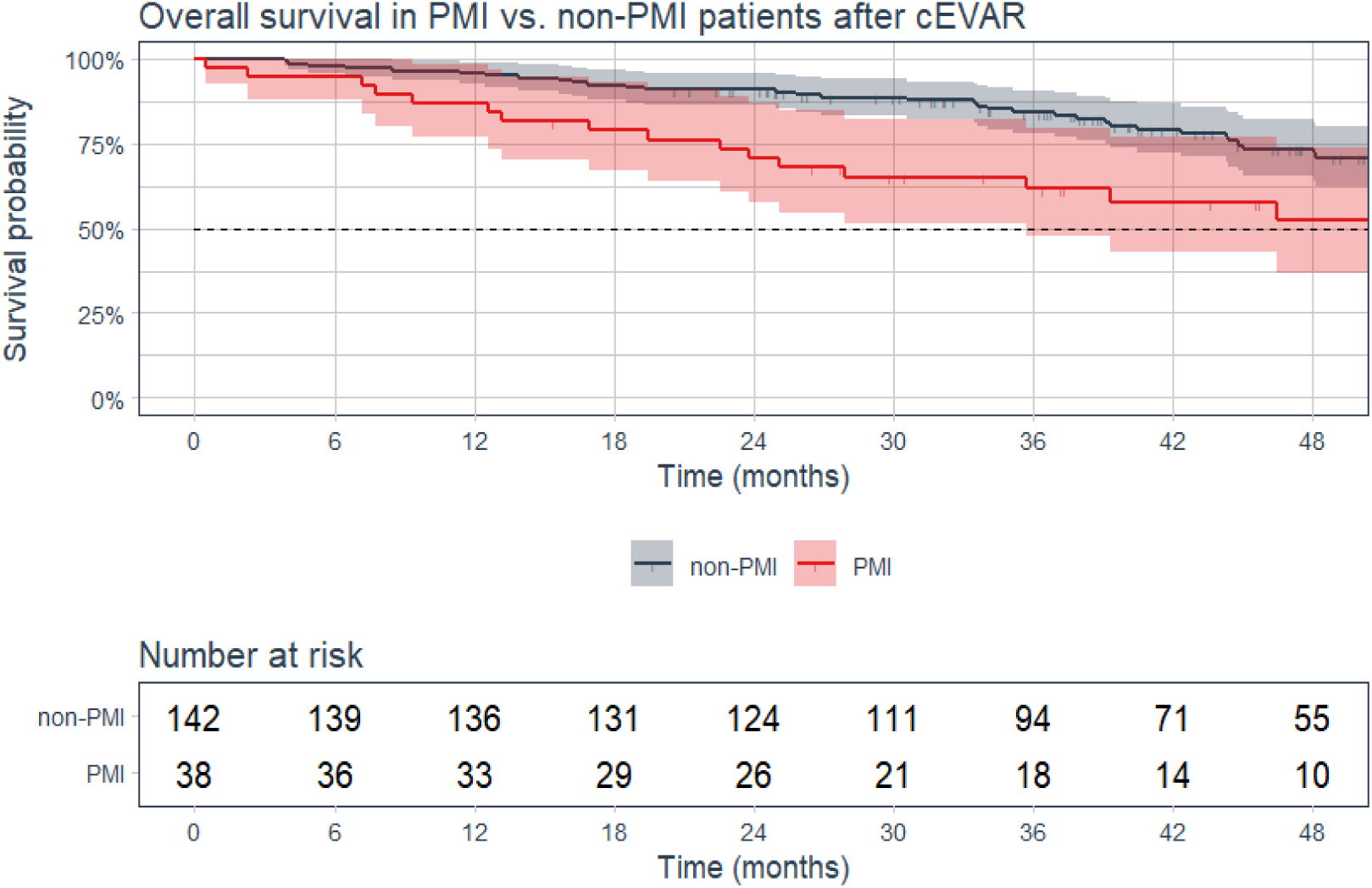
Cumulative Kaplan-Meier estimate of overall survival in patients with perioperative myocardial injury (PMI) compared to patients without PMI after complex endovascular aneurysm repair (c-EVAR).

PMI occurred in 38 of 180 patients (21.1%). Preoperative hsTnT levels exceeded the 99^th^ (>14 ng/L) percentile more frequently in patients with PMI compared to patients without PMI (68.4% vs. 45.8%) (Table 1). Median preoperative hsTnT levels in patients with PMI were also significantly higher compared to patients without (p = .019) (Table 2).

**Table 1:** comparison of baseline characteristics between patients with and without PMI after c-EVAR. Data are presented as n (%) or median (interquartile range 1, interquartile range 3). Missing values are excluded. ASA = American Society of Anesthesiologists; COPD = chronic obstructive pulmonary disease; DAPT = dual antiplatelet therapy; eGFR = estimated glomerular filtration rate; hsTnT = high sensitivity troponin T; PMI = peri-operative myocardial injury. * Patients with eGFR < 60 mL/min/1.73 m2 and no dialysis.

| Baseline characteristics | non-PMI<br>(N=142) | PMI<br>(N=38) | P-value |
| --- | --- | --- | --- |
| **Sex** |  |  | >.99 |
| Male | 114 (80.3) | 31 (81.6) |  |
| Female | 28 (19.7) | 7 (18.4) |  |
| **Age Mean (SD)** | 74.8 (5.37) | 75.5 (6.83) | .50 |
| **Smoking** |  |  | .62 |
| Non-smoker | 11 (7.8) | 3 (7.9) |  |
| Current smoker | 35 (24.6) | 11 (28.9) |  |
| Former smoker | 96 (67.6) | 23 (60.5) |  |
| **Hypertension** | 125 (88.0) | 31 (81.6) | .53 |
| **Ischaemic coronary disease** | 60 (42.3) | 26 (68.4) | **.012** |
| **Preoperative elevated hsTnT** | 65 (45.8) | 26 (68.4) | **.022** |
| **Peripheral arterial disease** | 45 (31.7) | 16 (42.1) | .31 |
| **COPD** | 47 (33.1) | 15 (39.5) | .59 |
| **Diabetes mellitus** | 17 (12.0) | 9 (23.7) | .12 |
| **Renal status** |  |  | .24 |
| eGFR: 15-60 ml/min | 65 (45.8) | 22 (57.9) |  |
| End stage CKD (eGFR<15) | 2 (2.1) | 0 (0) |  |
| Dialysis | 1 (0.7) | 0 (0) |  |
| **Statin** | 121 (85.2) | 30 (78.9) | .49 |
| **Beta Blocker** | 74 (52.1) | 20 (52.6) | >.99 |
| **Anti-platelet** |  |  | .56 |
| No anti-platelet | 45 (31.7) | 9 (23.7) |  |
| Aspirin | 96 (67.6) | 27 (71.1) |  |
| P2Y12 inhibitor | 1 (0.7) | 1 (2.6) |  |
| DAPT | 0 (0.0) | 1 (2.6) |  |
| **Anti-coagulation** | 39 (27.5) | 12 (31.6) | .77 |
| **Symptomatic aneurysm** | 2 (1.4) | 0 (0) | .46 |
| **Anemia** | 56 (39.4) | 19 (50.0) | .32 |
| **F/BEVAR** |  |  | .16 |
| Fevar | 94 (66.2) | 28 (73.7) |  |
| Bevar | 48 (33.8) | 10 (26.3) |  |

**Table 2:**
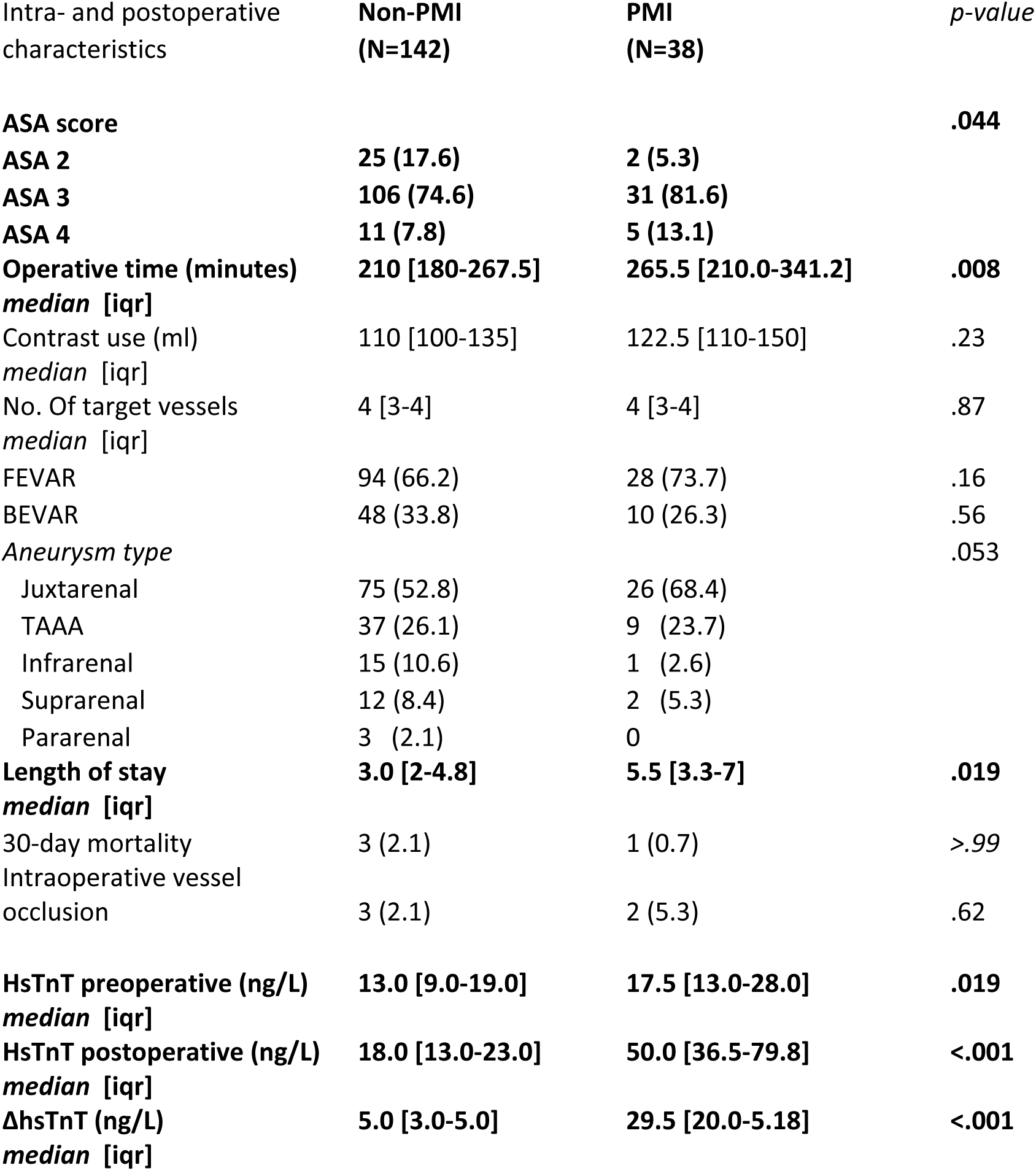
Comparison of in-hospital outcomes between patients with and without PMI after c-EVAR. Data are presented as n (%) or median (interquartile range 1, interquartile range 3). ASA = American Society of Anesthesiologists; eGFR = estimated glomerular filtration rate; hsTnT = high sensitivity troponin T; TAAA = thoraco abdominal aneurysm; F/BEVAR = fenestrated/branched Endovascular Aneursysm repair; PMI = peri-operative myocardial injury.

Patients with PMI significantly more often had a history of coronary artery (68.4% vs. 42.3%, p=.012). Furthermore, operative time and hospital length of stay were on average longer, and ASA scores were higher in these patients (Table 2).

The median follow-up time was 41 months [IQR: 28–57 months]. At 48 months, all-cause mortality was significantly higher in patients with PMI (42.1%, 16/38) compared to those without (21.8%, 31/142; p=.020). Patients with PMI had a 2.37-fold increased risk of four-year all-cause mortality following c-EVAR (hazard ratio (HR): 2.37 (95% CI: 1.29-4.33)). This association remained consistent after adjusting for age and sex (adjusted HR: 2.33 (95% CI: 1.27-4.27)), and persisted when additionally adjusting for a history of CAD, renal dysfunction, preoperative elevated hsTnT-levels and ASA score (adjusted HR: 2.37 (95% CI: 1.25-4.48)) (Table 3).

**Table 3:**
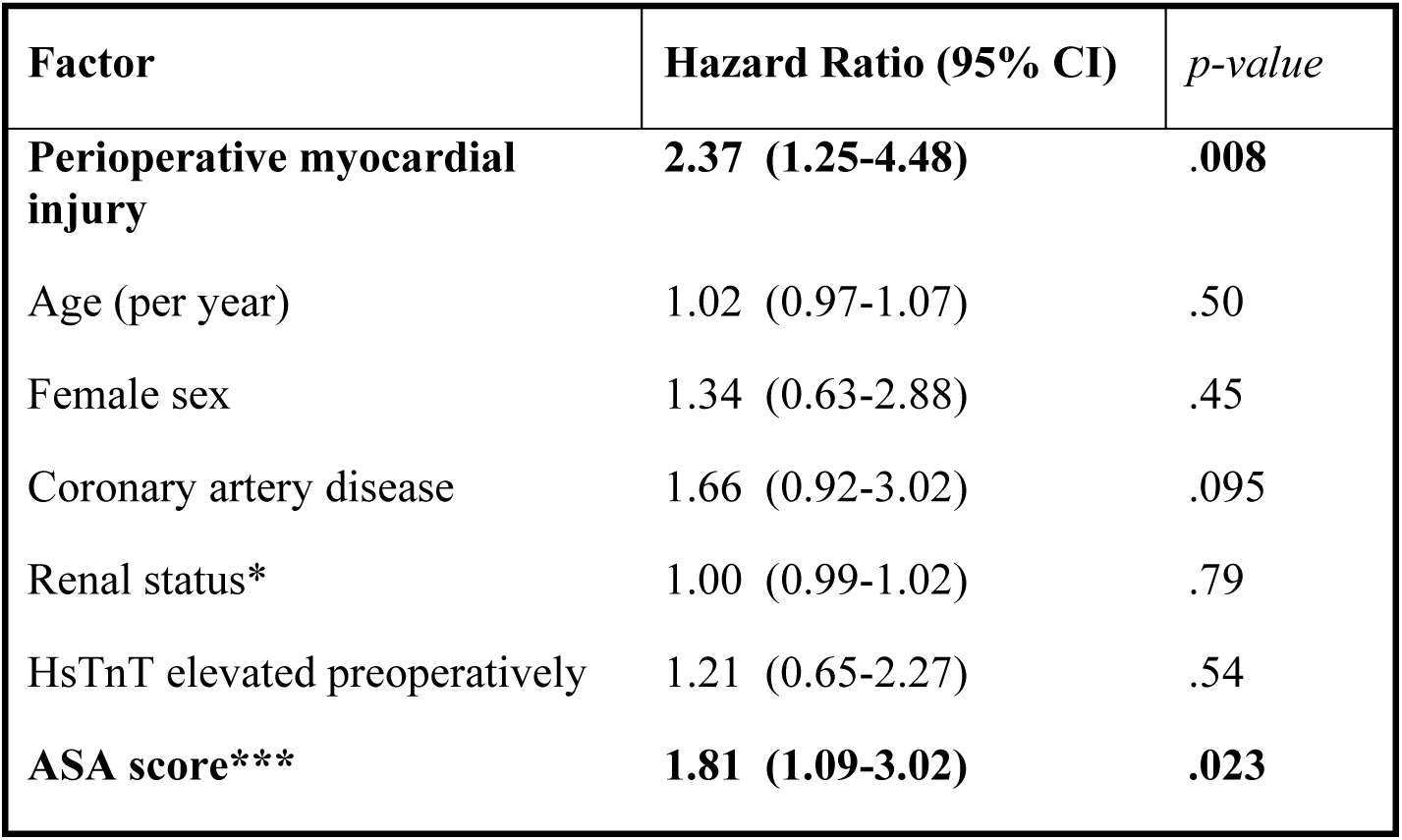
Multivariable Cox proportional hazards model for associative factors of mortality following c-EVAR. *patients with eGFR < 60 mL/min/1.73 m^2^ and no dialysis. ***Hazard ratio for every step up in ASA score. The reference is an ASA score of 2.

| Factor | Hazard Ratio (95% CI) | p-value |
| --- | --- | --- |
| **Perioperative myocardial injury** | **2.37 (1.25-4.48)** | **.008** |
| Age (per year) | 1.02 (0.97-1.07) | .50 |
| Female sex | 1.34 (0.63-2.88) | .45 |
| Coronary artery disease | 1.66 (0.92-3.02) | .095 |
| Renal status* | 1.00 (0.99-1.02) | .79 |
| HsTnT elevated preoperatively | 1.21 (0.65-2.27) | .54 |
| **ASA score\*\*\*** | **1.81 (1.09-3.02)** | **.023** |

**Table 4:**
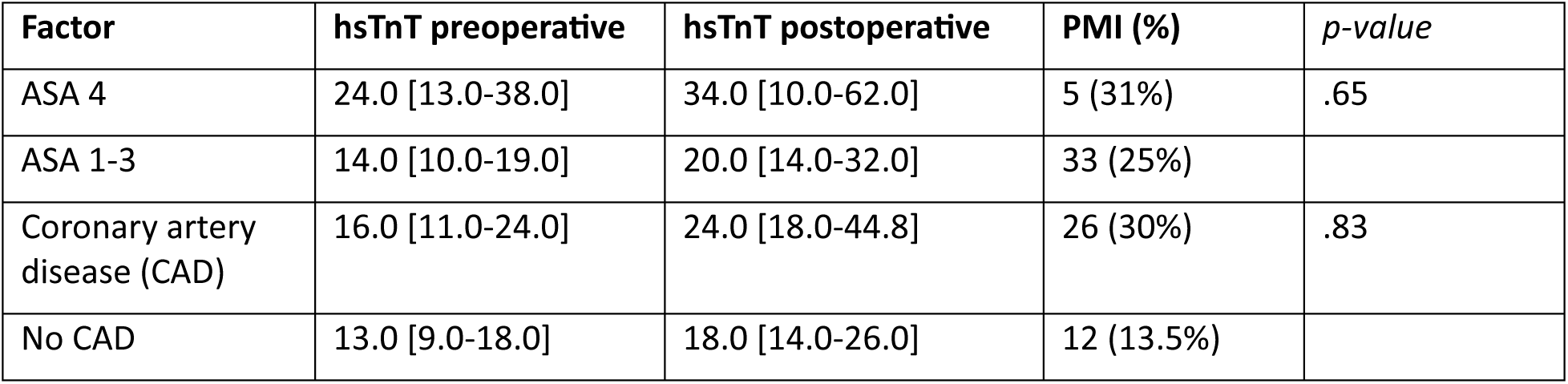
Median preoperative and postoperative hsTnT-levels (ng/L) in subgroups with baseline characterics more prevalent among patients with PMI (see Table 1). P-values reflect comparisons of postoperative hsTnT-levels between ASA-4 vs. ASA 1-3, eGFR <60 mL/min vs. eGFR >60 mL/min, and statin vs. no statin therapy. Values are presented as median [IQR].

| Factor | hsTnT preoperative | hsTnT postoperative | PMI (%) | p-value |
| --- | --- | --- | --- | --- |
| ASA 4 | 24.0 [13.0-38.0] | 34.0 [10.0-62.0] | 5 (31%) | .65 |
| ASA 1-3 | 14.0 [10.0-19.0] | 20.0 [14.0-32.0] | 33 (25%) |  |
| Coronary artery disease (CAD) | 16.0 [11.0-24.0] | 24.0 [18.0-44.8] | 26 (30%) | .83 |
| No CAD | 13.0 [9.0-18.0] | 18.0 [14.0-26.0] | 12 (13.5%) |  |

**Table 5:** multivariable logistic regression for associative risk factors for PMI following c-EVAR. *patients with eGFR < 60 mL/min/1.73 m^2^ and no dialysis. **OR per 1 ng/L increase in preoperative hsTnT with a maximum of preoperative hsTnT of 15 ng/L. ***Hazard ratio for every step up in ASA score. The reference is an ASA score of 2.

| Factor | Odds Ratio (95% CI) | p-value |
| --- | --- | --- |
| Age (per year) | 1.00 (0.94-1.06) | >.99 |
| Female sex | 1.55 (0.52-4.31) | .41 |
| Coronary artery disease | 2.38 (1.07-5.51) | .037 |
| Renal status* | 1.00 (0.97-1.02) | .99 |
| Preoperative HsTnT** | 1.11 (0.59-2.01) | .023 |
| ASA score*** | 2.95 (0.46-25.5) | .27 |

A history of coronary artery disease was significantly associated with an increased risk of developing PMI (adjusted OR: 2.38 (95% CI: 1.07-5.51)). Furthermore, preoperative elevated hsTnT-levels were independently associated with PMI (P=.023). Higher preoperative HsTnT-levels up to 15 ng/L were associated with an increased risk of developing PMI (OR: 1.22/Δ 1 ng/L, 95% CI: 1.04-1.47). Beyond 15 ng/L, higher preoperative hsTnT-levels did not further increase the risk of developing PMI (p>.05)(Figure 4).

**Figure 4:**
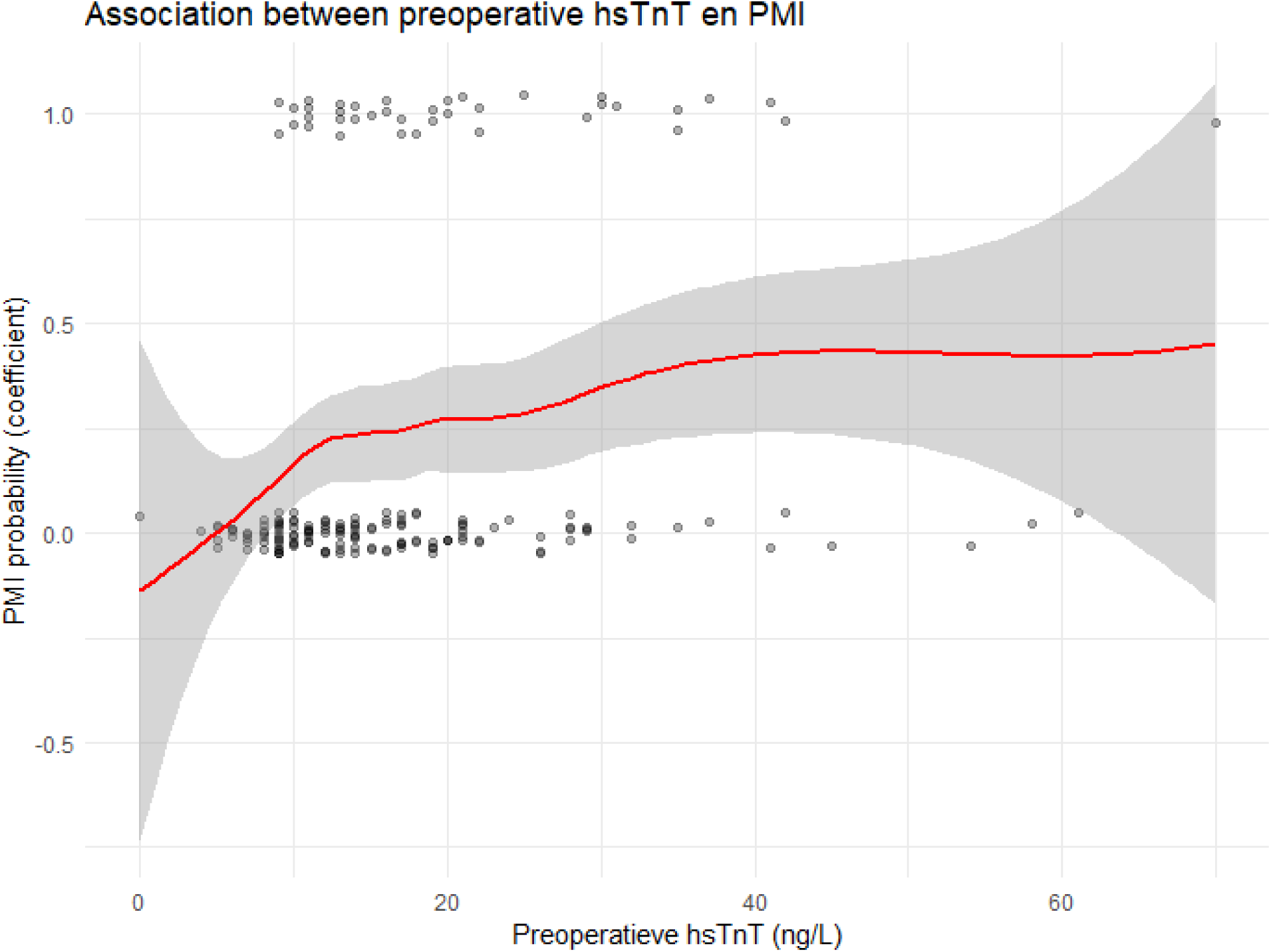
Association between individual preoperative hsTnT-levels (ng/L) and risk of developing perioperative myocardial injury (PMI) following c-EVAR visualized as survival probability for different degrees of preoperative hsTnT-levels.

## Discussion

Asymptomatic PMI is a relatively common event following complex EVAR. Using hsTnT, a sensitive marker to identify cardiac damage, PMI was found in approximately one out of five patients undergoing c-EVAR. Moreover, PMI following c-EVAR was associated with a more than twofold increased risk of mortality within four years, despite its asymptomatic character Studies about PMI in specific patient populations are scarce and uniform criteria regarding the minimum pre- to postoperative change in hsTnT-levels lack. However, our definition of PMI is in line with the latest recommendations.(1,2,12). To the best of our knowledge, our study is the first to consider a ΔhsTnT ≥ 14 ng/L in specifically c-EVAR, whereas other studies analyzing PMI following c-EVAR adhere to a minimum postoperative hsTnT value of ≥ 14 ng/L only.(13) Analyzing the increase between preoperative and postoperative troponin levels may better reflect cardiac strain associated with c-EVAR.

Our selection criteria for PMI yielded an incidence rate of 21%, which is considerably higher compared to infrarenal EVAR or non-cardiac surgery. Earlier studies about PMI in non-cardiac surgery in general found incidences ranging between 9% to 18%.(2,12) Similarly, PMI after infrarenal EVAR occurs in about 12% of patients.(14) The risk of death associated with PMI in c-EVAR was comparable to the long-term mortality risk of patients with PMI following infrarenal EVAR.(14)

In terms of 30-day mortality our results differ from previous studies which imply a twofold increase in 30-day mortality in patients who developed PMI, whereas our findings did not show any significant difference in 30-day mortality.(4,15) However, these studies primarily address non-cardiac surgery, including emergency interventions, whereas our study specifically focusses on elective c-EVAR.

Complex EVAR-patients, despite the procedure’s presumed minimally invasive character, seem to be specifically at risk to develop PMI, but the underlying mechanism remains unknown. PMI is believed to be a manifestation of a sudden mismatch in cardiac oxygen supply and demand, potentially caused by underlying cardiovascular comorbidities. Almost three-quarters of the patients with PMI had troponin levels that were already elevated preoperatively, suggesting there is an underlying condition causing a chronic mismatch in cardiac oxygen supply and demand. Correspondingly, about half of the patients with PMI had a history of CAD, or coronary revascularisation, while this was only the case of about one fourth of the patients without PMI. Interestingly, earlier studies suggested that up to 60% of patients with (complex)-AAAs suffer from occult CAD, i.e. underdiagnosed or undertreated CAD due to the absence of symptoms.(16,17) In daily activities or routine exercise cardiac oxygen supply may still be sufficient.(13) However, prolonged or increased cardiac stress still results in silent myocardial ischaemia. In the case of c-EVAR-patients, myocardial ischaemia could be the result of cardiac strain caused by the intervention and anaesthesiology, potentially leading to an acute rise in troponin levels and subsequent PMI.(18,19) In this context, the c-EVAR procedure could function akin to a cardiac stress test, identifying patients who may be at increased cardiac risk.

Contradictory, in our study not all patients with proven CAD developed PMI (PMI occurred in 34%). Furthermore, 55% of the patients with proven CAD had preoperative elevated troponin levels, but levels remained stable throughout their hospital stay in half of these patients. Why some patients with a history of CAD seem to be more susceptible to PMI should be a topic for future studies. Nevertheless, our results suggest that PMI (i.e. an acute rise in troponin levels) is a stronger predictor for postoperative mortality than preoperative troponin levels alone. Preoperative hsTnT levels up to 15 ng/L were associated with an increasing risk of developing PMI, whereas this risk plateaued at levels above 15 ng/L. Moreover, in contrast to PMI there was no significant association between preoperative hsTnT-levels and mortality. This supports the clinical relevance of measuring both preoperative and postoperative troponin levels and the uniformly accepted cutoff value of hsTnT at 14 ng/L, consistent with previous studies.(12,18,20)

Furthermore, ASA scores tend to be higher in patients with PMI, indicating an increased overall vulnerability among those who subsequently develop PMI. Also, our results suggest that operative time and length of hospital stay were longer in patients with PMI, which is also substantiated in other studies (21,22). Hypotension, hypothermia, tachycardia, and inflammation are other possible perioperative contributors to PMI and require further investigation. (23,24) This leaves the impression PMI reflects a multifactorial process in which, next to CAD, perioperative factors, such as hemodynamic fluctuations, anatomical complexity, and patient frailty in general also play a role.(22)

Finally, further investigation is warranted to assess the impact of optimizing cardiovascular medication on long-term mortality in PMI-patients. Although in this study no significant association was found between medication treatments and mortality, our prior research on PMI following infrarenal EVAR demonstrated a protective effect of statin therapy on long term mortality.(14) The benefits of statin, such as cholesterol reduction and pleiotropic effects, have also been documented in other studies.(25) Despite this, patients are often discharged with insufficient cardiovascular medication regimens, which may contribute to increased susceptibility to PMI and higher long-term mortality.(20)

### Limitations

Given the limited number of events, covariates were limited to the covariates above and other factors were not included in order to obtain unbiased estimates with certainty.(26) Although several baseline characteristics that seem to occur more frequently in patients with PMI compared to patients without, the limited sample size and single-center set up prevented further statistical analyses to investigate the association of these characteristics with PMI. Furthermore, due to the study’s retrospective character, only variables already included in our database were used for analysis. Larger, multicenter prospective studies should be aimed for to incorporate more confounding variables including coronary imaging assessments and determine their association with PMI following c-EVAR. Nevertheless, various statistical tests were applied to assess the association between PMI and mid-term mortality, which resulted in a robust outcome. Therefore, we consider to have appropirately identified PMI as significant predictor for mid-term mortality.

The potential impact of acute kidney injury (AKI) also requires further analyses as impaired renal function may contribute to higher circulating troponin levels.(27) Although our analyses adjusted for baseline renal function, reliable data on perioperative changes in renal function was unavailable due to the study’s retrospective character. In particular, the potential association between intraoperative complications, such as renal artery occlusion, AKI and PMI, should be evaluated in larger cohorts with more events of PMI.

Due to the retrospective character of this study, routine invasive cardiac screening for AMI in all postoperative patients was not possible. Consequently, some overestimation of PMI at the expense of AMI cannot be excluded, since clinical signs of myocardial ischaemia may be masked by postoperative analgesia.(28) Nonetheless, approximately two-thirds of the patients underwent postoperative cardiac assessment within 72 hours after surgery, typically due to clinical suspicion of myocardial ischaemia, intensive care monitoring protocols, or enrolment in anaesthesiology related studies. AMI was diagnosed in two patients through this process, both of whom were excluded from this study’s analyses. Given that a substantial proportion of the cohort underwent targeted postoperative screening, we consider the estimated incidence of PMI to be reliable. Furthermore, comprehensive documentation regarding the definitive causes of death in PMI-patients was unavailable. Prospective studies are warranted to accurately capture mortality attributed to cardiac events.

### Clinical implications

Despite the limited number of events, all three Cox-regressions yielded robust results, revealing a significant association between asymptomatic/subclinical PMI and mortality. Therefore, we concur that PMI following c-EVAR has been justifiably identified as relevant predictor for mid-term mortality. Routine perioperative hsTnT monitoring in c-EVAR-patients could contribute to the identification of at-risk patients. Given the high prevalence of cardiac comorbidities, such as CAD, c-EVAR-patients with perioperative elevated hsTnT-levels could benefit from intensified postoperative cardiac screening and possible (invasive) therapy. Future studies should clarify whether improved postoperative cardiac management and treatment could result in improved long-term survival.

## Data Availability

The data that support the findings of this study are not publicly available due to privacy restrictions, but anonymized data may be available from the corresponding author upon reasonable request and with approval from the Erasmus University Medical Center.

